# Persistence of immunity and impact of a third (booster) dose of an inactivated SARS-CoV-2 vaccine, BBV152; a phase 2, double-blind, randomised controlled trial

**DOI:** 10.1101/2022.01.05.22268777

**Authors:** Krishna Mohan Vadrevu, Brunda Ganneru, Siddharth Reddy, Harsh Jogdand, Dugyala Raju, Usha Praturi, Gajanan Sapkal, Pragya Yadav, Prabhakar Reddy, Savita Verma, Chandramani Singh, Sagar Vivek Redkar, Chandra Sekhar Gillurkar, Jitendra Singh Kushwaha, Satyajit Mohapatra, Amit Bhate, Sanjay Rai, Raches Ella, Priya Abraham, Sai Prasad, Krishna Ella

## Abstract

**Background:** Neutralising antibody responses to SARS-CoV-2 vaccines have been reported to decline within 6 months of vaccination, particularly against Variants of Concern (VOC). We assessed the immunogenicity and safety of a booster dose of BBV152 administered 6 months after the second of a two-dose primary vaccination series.

**Methods:** In an ongoing phase 2 trial (ClinicalTrials.gov: NCT04471519) the protocol was amended after six months to re-consent and randomise 184 previously vaccinated participants to receive a third dose of vaccine or placebo on Day 215. The primary outcome was to measure neutralising antibody titres by plaque-reduction neutralisation test (PRNT_50_) four weeks after the booster; safety as serious adverse events (SAE) was the key secondary outcome.

**Findings:** Four weeks after a second BBV152 vaccination geometric mean titres (GMTs) of neutralising antibodies were 197·0 PRNT_50_ (95% CI: 155·6–249·4); this level declined to 23·9 PRNT_50_ (14·0–40·6) six months later, with a seroconversion rate of 75·4% (95% CI: 68·4–81·6). Four weeks after booster vaccination the GMT increased on Day 243 to 746·6 PRNT_50_ (514·9–1081) compared with 100·7 PRNT_50_ (43·6–232·6) in the placebo group. Corresponding seroconversion rates were 98·7% (92·8–99·9) and 79·8% (69·6–87·8). Increased titres in the placebo group were attributed to natural infection as the study was conducted during the second wave of COVID-19 in India. PRNT_50_ titres against the SARS-CoV-2 variants increased—Alpha (32·6-fold), Beta (161·0-fold), Delta (264·7-fold), and Delta plus (174·2-fold)—after the booster vaccination. We found that vaccine induces both memory B and T cells with a distinct AIM+ specific CD4+T central and effector memory phenotype, including CD8+ T_EMRA_ phenotype. Reactogenicity after vaccine and placebo was minimal and comparable, and no SAEs were reported.

**Interpretation:** Six months after a two-dose BBV152 vaccination series cell mediated immunity and neutralising antibodies to both homologous (D614G) and heterologous strains (Alpha, Beta, Delta and Delta plus) persisted above baseline, although the magnitude of the responses had declined. Neutralising antibodies against homologous and heterologous SARS-CoV-2 variants increased 19- to 97-fold after a third vaccination. Booster BBV152 vaccination is safe and may be necessary to ensure persistent immunity to prevent breakthrough infections.

**Funding:** This work was supported and funded by Bharat Biotech International Limited.

## Introduction

The emergence of SARS-CoV-2 variants of concern (VoC) has raised concerns about the breadth and durability of neutralising antibody responses [1]. Diminished vaccine effectiveness against the Beta (B.1.351) and Delta (B.1.617.2) VoCs has been reported for several authorised vaccines [2–5]. With the potential of newly emergent highly transmissible VoCs, illustrated by the recent circulation of the Omicron variant [6], understanding the persistence of neutralising antibody responses to convery long-term efficacy is vital to assess the need for additional booster doses.

BBV152 is a whole-virion inactivated SARS-CoV-2 vaccine formulated with a Toll-like receptor 7/8 agonist molecule adsorbed onto alum (Algel-IMDG). We previously reported interim findings from a phase 2 controlled, randomised, double-blind trial on the immunogenicity and safety of two different formulations of BBV152 intended to select one formulation for further clinical development [7]. Based on tolerable safety outcomes, humoral and cell-mediated responses the 6 µg dose with Algel-IMDG was selected for assessment in a phase 3 efficacy trial in which we demonstarted an overall vaccine efficacy of 77·8% (95% CI: 65·2–86·4) against any COVID-19 and 65·2% (95% CI: 33·1–83·0) efficacy against the Delta (B.1.617.2) VoC [8].

Trial participants in the phase 2 trial (hereafter referred to as the parent study) were followed up until 6 months after the second dose to evaluate the durability of immune responses. Following a protocol amendment and obtention of new consent we randomised participants who previously received the 6 µg dose with Algel-IMDG to receive either a third (booster) dose of BBV152 or placebo. We report our findings on the immunogenicity and safety of that third (booster) dose of BBV152.

## Methods

### Trial Design and Participants

The parent study was a randomised, double-blind, multicentre phase 2 trial to evaluate the immunogenicity and safety of a whole-virion inactivated SARS-CoV-2 vaccine (BBV152) in healthy male and female volunteers in nine Indian hospitals [7]. The trial was approved by the National Regulatory Authority (India) and the respective hospital Ethics Committees and conducted in compliance with all International Council for Harmonization (ICH) Good Clinical Practice guidelines. The trial protocol was registered on ClinicalTrials.gov: NCT04471519. Participants were ≥ 12 to 64 years of age at the time of enrolment, and were negative for both SARS-CoV-2 nucleic acid and serology tests at baseline (before receiving the primary vaccination series).

In the parent study a total of 380 participants had been randomised 1:1 to receive BBV152 formulations containing either 3 µg or 6 µg doses of antigen with Algel-IMDG and were followed up for 6 months after dose 2 for persistence of safety and immunogenicity. Additional details of the parent study have previously been reported [7]. Following an amendment to the protocol 6 months after dose 2, new informed consent was obtained from participants who originally received the 6 µg dose of BBV152, including some who had “dropped out” of the parent study. These participants were then randomised 1:1 to receive either a third (booster) dose of vaccine or placebo. Using a block size of four, randomisation was done by the unblinded contract research organisation (CRO) managing the study using a master randomisation code uploaded to an Interactive Web Response System (Sclin Soft Technologies). A central depot manager was responsible for uploading the vials containing a unique code to the respective sites, where a masked study nurse was responsible for vaccine preparation and administration.

Participants, investigators, study coordinators, study-related personnel, and the sponsor masked the treatment group allocation.

### Trial Vaccine

BBV152 (manufactured by Bharat Biotech) is a whole-virion ß-propiolactone-inactivated SARS-CoV-2 vaccine adjuvanted with Algel-IMDG. The virus strain NIV-2020-770 was isolated from a COVID-19 patient, sequenced at the Indian Council of Medical Research-National Institute of Virology (NIV), and provided to Bharat Biotech [9]. The vaccine virus strain NIV-2020-770 contains the D614G mutation, characterised by an aspartic acid to glycine shift at amino acid position 614 of the spike protein [9]. The control arm received a placebo containing sterile phosphate-buffered saline and Algel. Vaccine and control formulations were supplied as 0·5mL doses in single-use glass vials, stored between 2°C and 8°C. The appearance, colour and viscosity, of vaccine and placebo, were identical. Vaccine and placebo were administered by intramuscular injection in the deltoid muscle.

### Outcomes

The primary endpoints were neutralising antibody titres against wild-type virus evaluated by two neutralisation assays; a plaque-reduction neutralisation test (PRNT) and a microneutralisation assay (MNT) done at Bharat Biotech laboratories (Hyderabad, India) as previously described [7–9]. Secondary endpoints were the percentages of participants with solicited local and systemic reactogenicity within seven days after vaccination. As exploratory endpoints, neutralising responses against SARS-CoV-2 variants and cell-mediated immune responses were evaluated.

### Procedures

All participants from the phase 2 parent study were followed up until Day 208 (6 months after dose 2) to evaluate the durability of the immune response. In the randomised cohort enrolled into the extension study, a blood sample was taken on Day 215, and then a vaccine or placebo was administered. Participants were observed for two hours post-vaccination to assess immediate reactogenicity. They were instructed to record solicited local reactions and systemic adverse events within seven days of the booster dose using a paper-based memory aid, which included fields for time of symptom onset, severity, time to resolution, and concomitant medications. No prophylactic medication (ibuprofen/acetaminophen) was prescribed before or after vaccination. Participants were instructed to complete the form daily, and it was collected during the next visit to the site. In addition, routine telephone calls were scheduled during the first seven days after vaccination.

Solicited local adverse events included pain at the injection site and swelling. Systemic adverse events included fever, fatigue/malaise, myalgia, body aches, headache, nausea/vomiting, anorexia, chills, generalised rash, and diarrhoea. Participants throughout the study reported any unsolicited adverse events. Adverse events were graded according to the severity score (mild, moderate, or severe) and whether they were related or unrelated to the investigational vaccine, as detailed in the protocol. At a follow-up visit on Day 243, safety data were collected, and a second blood sample was drawn for immunogenicity assessments.

### Immunogenicity

Neutralising antibody titres against wild type virus was measured by live virus neutralisation at Bharat Biotech, Hyderabad [7–9] and a subgroup of serum samples were assessed for neutralising antibody titres against heterologous strains by plaque-reduction neutralisation test (PRNT_50_) at the National Institute of Virology (NIV, Pune, India). IgG-binding antibody binding titres were determined by ELISA against the SARS-CoV-2 specific proteins: Spike protein (S1), receptor-binding domain (RBD) and nucleocapsid protein (N). As there is no established SARS-CoV-2 correlate of protection, vaccine-induced responses were compared with an internationally recognised reference serum (BEI, Biodefense and Emerging Infections Research Resources Repository, NIAID, NIH, USA).

Peripheral blood mononuclear cells (PBMC) were collected from a subset of participants randomly selected using a code from the CRO and assessed SARS-CoV-2-specific T and B cell memory responses on Days 215 243 in a blinded manner at Immunitas Biosciences (Bengaluru, India). T cell responses were assessed by activation-induced marker (AIM) in PBMC stimulated with overlapping peptide pools of SARS-CoV-2 S, M, and N proteins. In the ELISpot assay, PBMCs were stimulated for polyclonal stimulation followed by inactivated SARS-CoV-2 antigen to evaluate antigen-specific memory B-cells (MBC). SARS-CoV-2-specific IFNγthe release was evaluated using a whole blood T-cell immunity assay from a subset of vaccinated participants on Day 208. Detailed assay methods are provided in the *Supplementary Appendix*, pages 2–3. Finally, we assessed for a Th1 biased immune response based on the ratio of IgG1 and IgG4 endpoint antibody titres in placebo and vaccine recipients on Days 215 and 243.

### Statistical Analysis

No formal sample size estimation was made to compare differences in neutralising antibody responses between the booster and control arm. Safety endpoints are presented descriptively as frequencies (%) per group. Immunological endpoints are presented as group GMTs with 95% confidence intervals (CIs) and group seroconversion rates; seroconversion was defined as post-vaccination titre ≥ 4-fold above the pre-vaccination titre in each participant. For continuous variables (below 20 observations), medians and IQRs are reported. The exact binomial calculation was used for the CI estimation of proportions. Wilson’s test was used to test differences in proportions. The GMT confidence intervals were estimated based on the log_10_ (titre) and the assumption that the log_10_ (titre) was normally distributed. A comparison of GMTs was performed with t-tests on the means of the log_10_ (titre). Significance was set at p < 0·05 (2-sided). Descriptive and inferential statistics were performed using SAS 9·2.

## RESULTS

Of the 190 participants who originally received the 6 µg BBV152 formulation with Algel-IMDG in the parent study between Sep 7 and Sep 11, 2020, 175 participants were still actively participating at Day 208, with 14 drop-outs (**Figure 1**). On being recontacted 9 of the 14 who had dropped out agreed to re-consent for the extension study. Thus, a total of 184 participants were re-enrolled on Day 215 and randomised 1:1 to receive either a booster dose of BBV152 or a placebo. Demographic characteristics of these participants are shown in **Table 1**.

**Figure 1.**
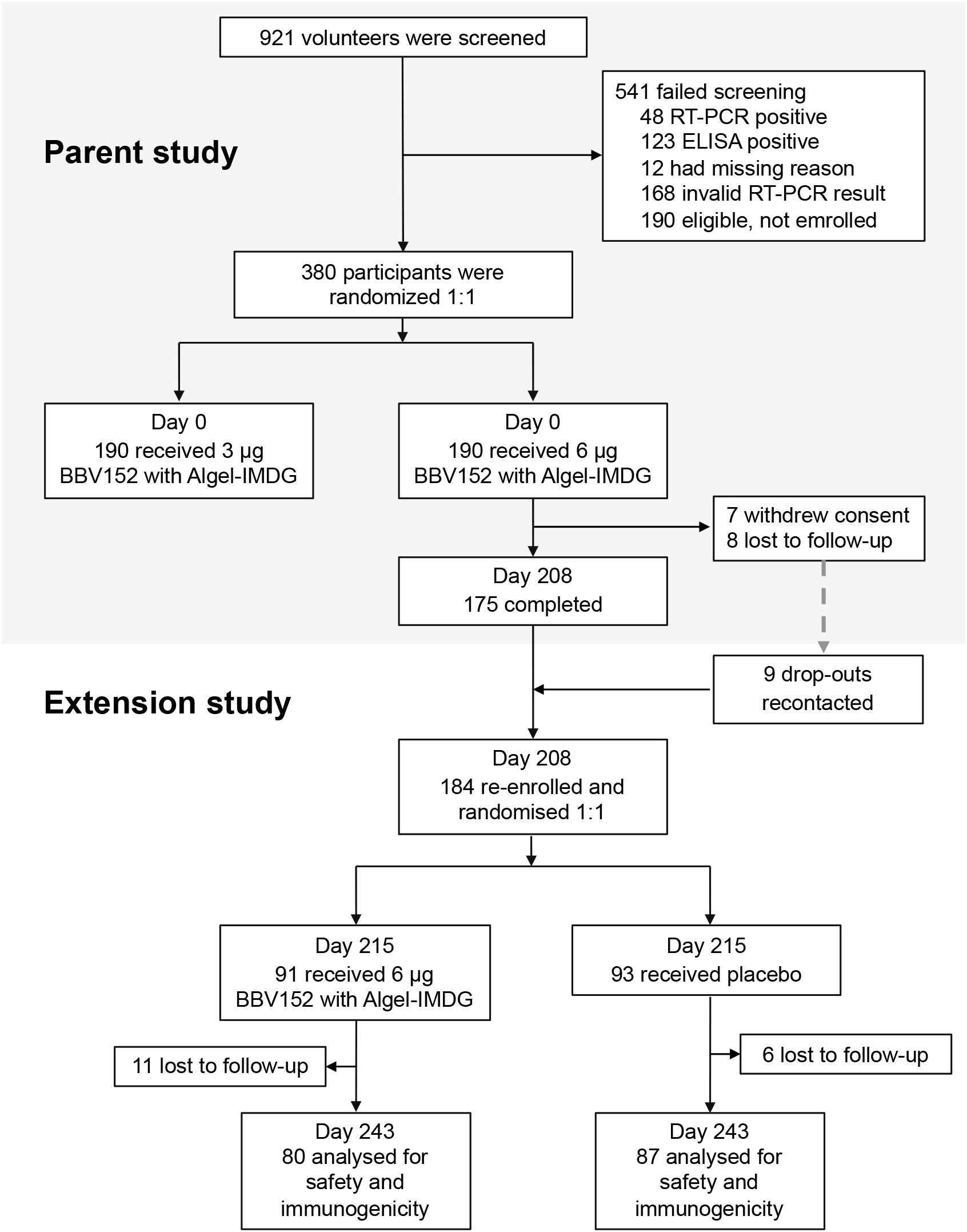
Study flow chart.

**Figure 2:**
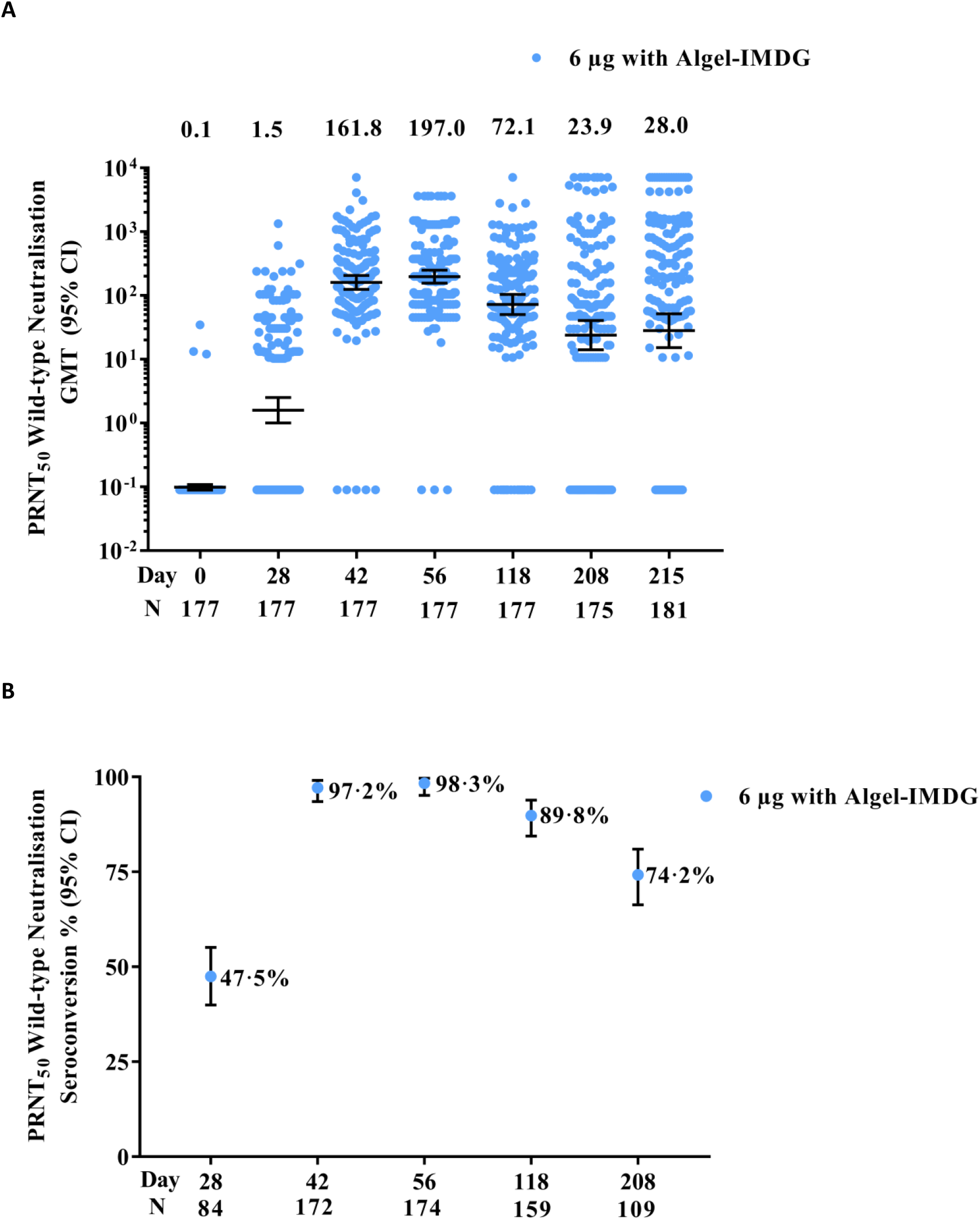
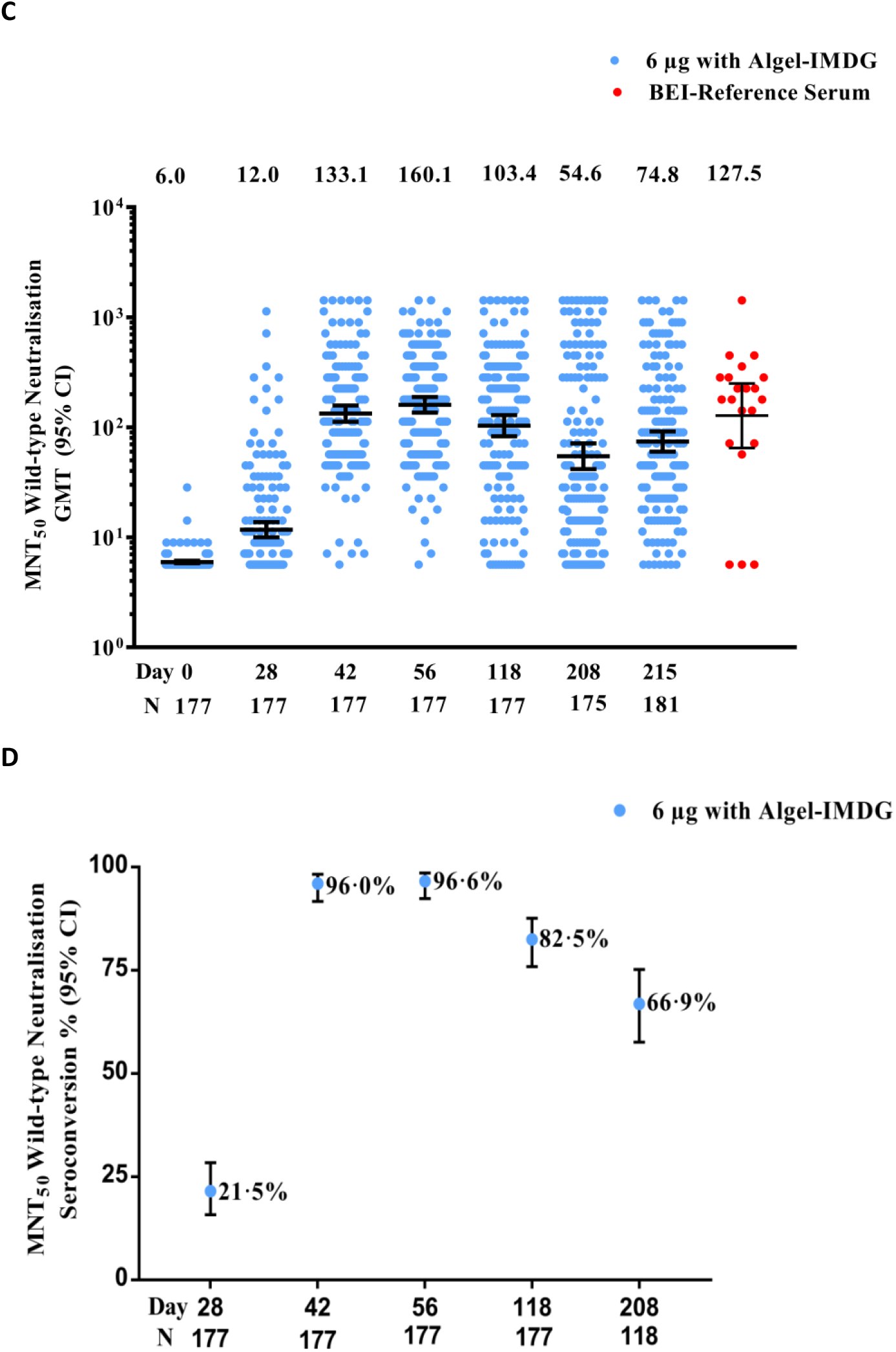
SARS-CoV-2 Live Virus Neutralizing Responses (post-primary immunization series) Titres of the wild-type SARS-CoV-2 neutralisation assay (PRNT50 and MNT50) at baseline (day 0), 4 weeks after the first vaccination (day 28), 2 weeks after the second vaccination (day 42), and 4 weeks after the second vaccination (day 56), 12 weeks after the second vaccination (day 118), 24 weeks (6 months) after the second vaccination (day 208), and 25 weeks after the second vaccination (day 215) for 6 µg with Algel-IMDG groups are shown. SCRs were defined based on the proportion of titres ≥4-fold above baseline. The dots and horizontal bars represent the SCR and 95% CI, respectively (panels A&C). In panels B&D, the dots and horizontal bars represent individual data points and the geometric mean (95% CI). The BEI Reference serum (BEI) panel was evaluated in MNT alone.

**Figure 3:**
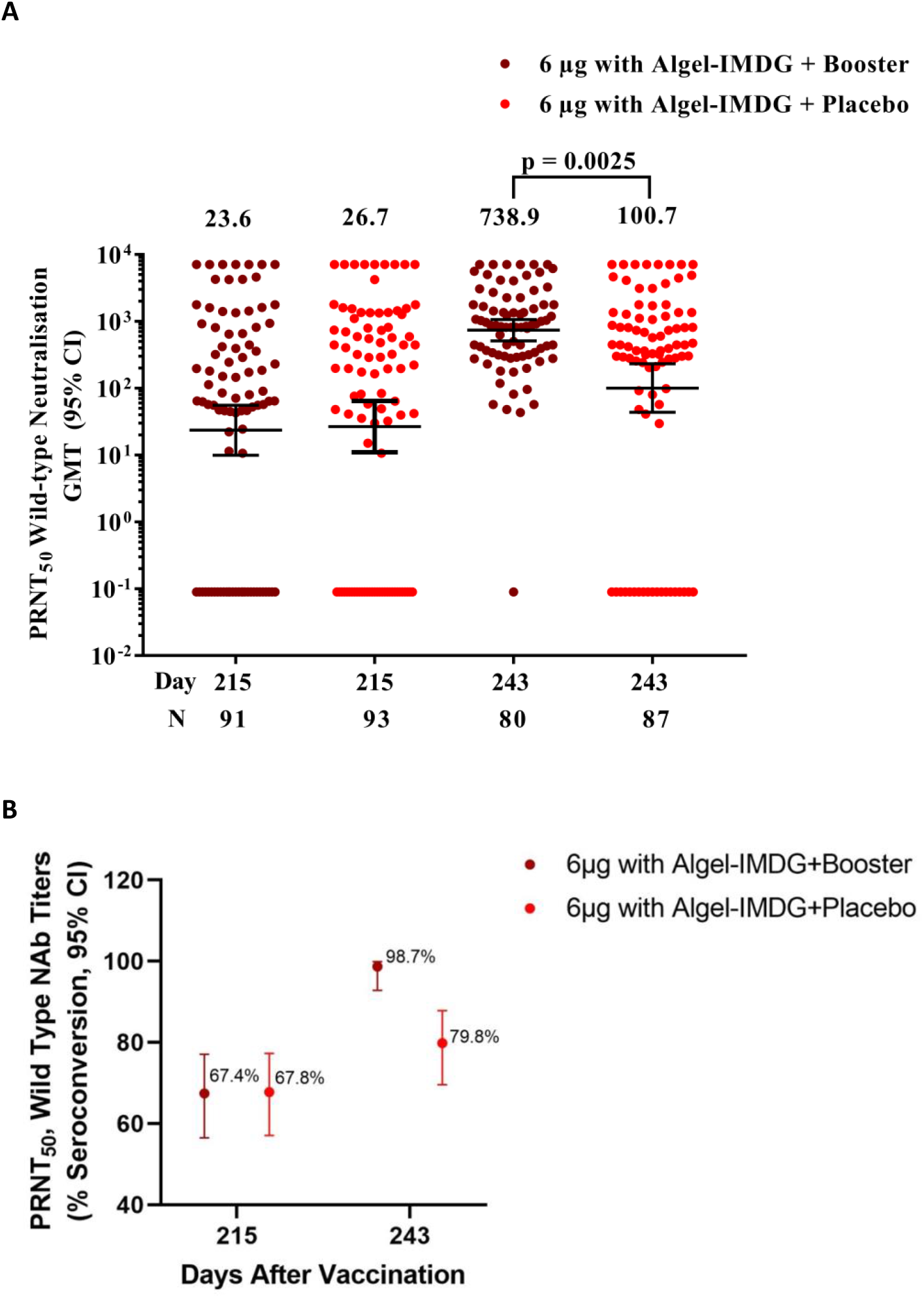
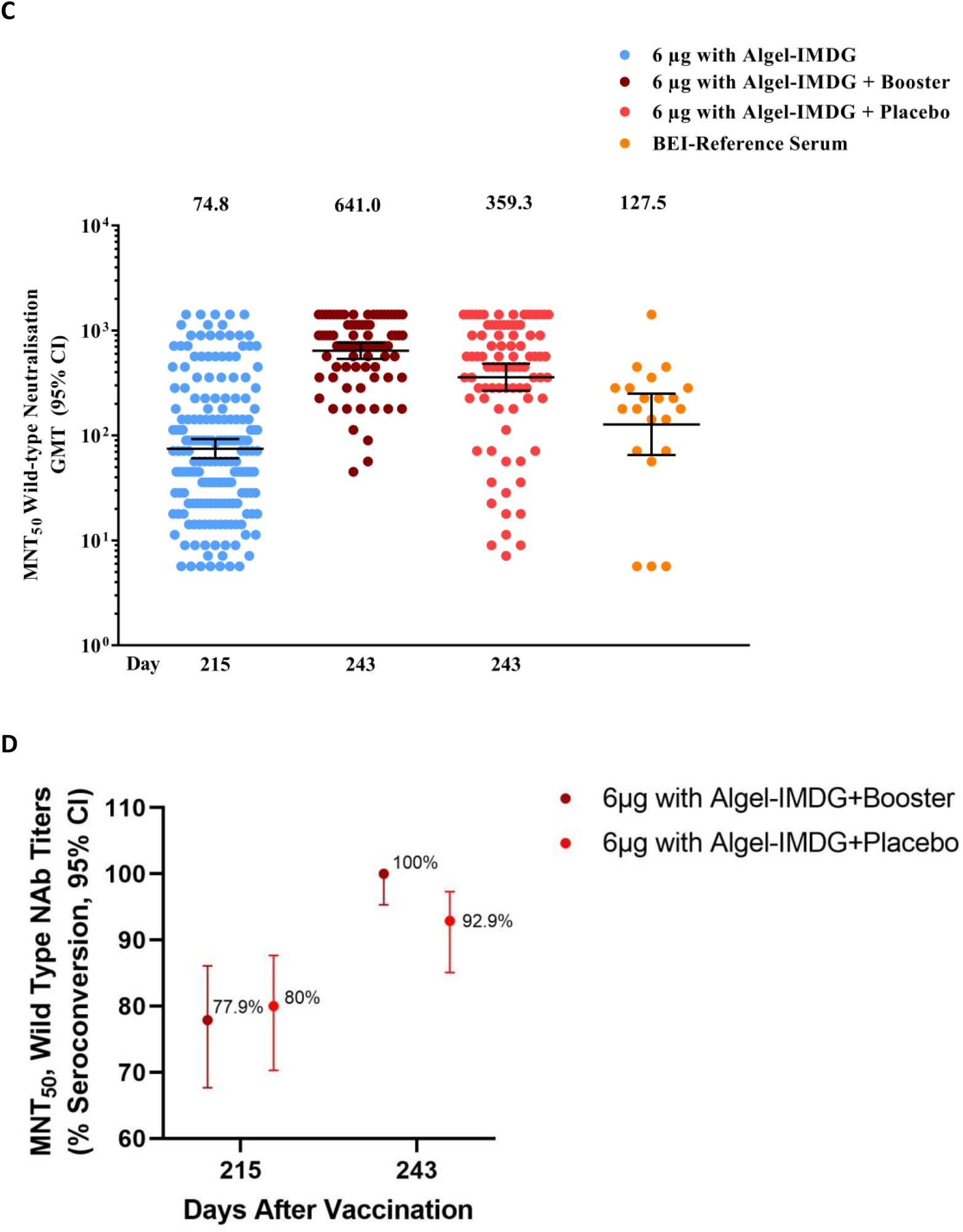
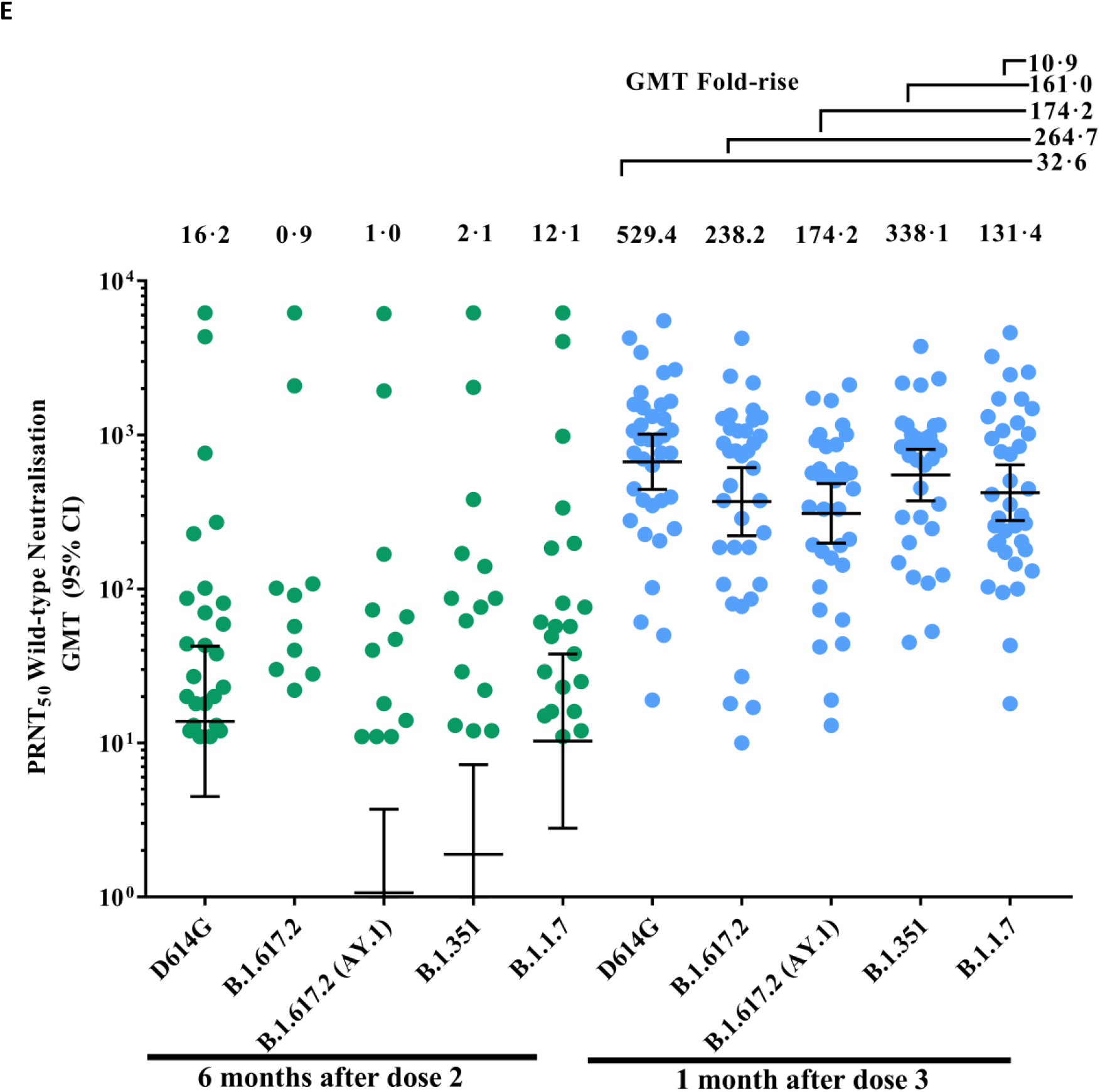
SARS-CoV-2 Live Virus Neutralizing Responses (post homologous booster dose) Titres of the wild-type SARS-CoV-2 neutralisation assay (PRNT_50_ and MNT_50_) at 25 weeks after the second vaccination (day 215) and 4 weeks after the third dose (booster) or placebo (day 243). SCRs were defined based on the proportion of titres ≥4-fold above baseline (day 0). In panels A&C, the dots and horizontal bars represent individual data points and the geometric mean (95% CI). The dots and horizontal bars represent the SCR and 95% CI, respectively (panels B&D). In panel E, for the booster dose arm only, the dots and horizontal bars represent individual data points, the geometric mean (95% CI), and geometric fold-rise (1 month after dose 3 Vs 6 months after dose 2).

**Table 1:** Demographics of participants enrolled in the booster dose study

|  |  |  |
| --- | --- | --- |
|  | **Vaccine (N = 91)** | **Placebo (N = 93)** |
| **Age, years** |  |  |
| Median | 35·0 (25·0–44·0) | 36·0 (26·0–44·0) |
| ≥ 12 to < 18 | 0 (0%) | 4 (4·3%) |
| ≥ 18 to < 55 | 85 (93·4%) | 82 (88·2%) |
| ≥ 55 to ≤ 65 | 6 (6·6%) | 7 (7·5%) |
| **Sex** |  |  |
| Female | 24 (26·4%) | 20 (21·5%) |
| Male | 67 (73·6%) | 73 (78·5%) |
| **Body Mass Index, kg/m<sup>2</sup>** | 25·3 (3·5) | 24·7 (2·7) |
| Data are median (IQR), n (%) or mean (SD) |  |  |

### Neutralising antibodies

As previously reported in the parent study two 6 µg doses of BBV152 administered 28 days apart induced high neutralising antibody titres [7]. Thus, on Day 56, four weeks after the second dose, the PRNT_50_ GMT was 197 (95% CI: 156–249), and the MNT_50_ GMT was 160 (136–189). Seroconversion was observed in 174 (98·3% [95% CI: 95·1–99·7]) of 177 participants by PRNT, and in 171 (96·6% [92·8–98·8]) of 177 participants by MNT (**Table 2**).

**Table 2:** Neutralising antibody titres against SARS-CoV-2 measured by PRNT and MNT.

|  |  | Geometric mean titre<br>(95% CI) |  | Seroconversion rate <sup>a</sup><br>(95% CI) |  |
| --- | --- | --- | --- | --- | --- |
|  |  | Booster | Placebo | Booster | Placebo |
| Plaque reduction neutralisation test (PRNT) |  |  |  |  |  |
| Parent study | Day 56 | N = 177 |  | 174/177 |  |
|  |  | 197.0<br>(155.6–249.4) |  | 98.3%<br>(95.1–99.6) |  |
| Booster study | Day 215 | N = 91 | N = 93 | N = 91 | N = 93 |
|  |  | 23.6<br>(10.0–55.7) | 26.7<br>(11.0–64.6) | 67.4<br>(56.5–77.1) | 67.8<br>(57.1–77.3) |
|  | Day 243 | N = 80 | N = 87 | N = 80 | N = 87 |
|  |  | 746.2<br>(514.9–1081)<br>*p* < 0.0001<sup>b</sup> | 100.7<br>(43.6–232.6)<br>*p* < 0.0269<sup>b</sup> | 98.7<br>(92.8–99.9) | 79.8<br>(69.6–87.8) |
| Microneutralisation test (MNT) |  |  |  |  |  |
| Parent study | Day 56 | N = 177 |  | 171/177 |  |
|  |  | 160.1<br>(135.8–188.8) |  | 96.6%<br>(92.8–98.8) |  |
| Booster study | Day 215 | N = 91 | N = 93 | N = 91 | N = 93 |
|  |  | 66.8<br>(49.7–89.7) | 85.0<br>(62.5–115.5) | 77.9<br>(67.7–86.1) | 80.0<br>(70.3–87.7) |
|  | Day 243 | N = 80 | N = 87 | N = 80 | N = 87 |
|  |  | 641.0<br>(536.8–765.3)<br>*p* < 0.0001<sup>b</sup> | 359.3<br>(267.4–482.7)<br>*p* < 0.0001<sup>b</sup> | 100<br>(95.3–100) | 92.9<br>(85.1–97.3) |
| PRNT GMTs against SARS-CoV-2 variants |  |  |  |  |  |
| Variant | D614G | Alpha | Beta | Delta | Delta plus |
| Day 208 | N = 91 | N = 91 | N = 91 | N = 91 | N = 91 |
|  | 33.3<br>(15.6–71.0) | 10.3<br>(3.0–35.8) | 2.9<br>(1.2–7.4) | 2.6<br>(1.0–6.8) | 1.9<br>(0.5–6.9) |
| Day 243 | N = 80 | N = 80 | N = 80 | N = 80 | N = 80 |
|  | 632.3<br>(380.2–1052) | 338.1<br>(188.2–607.4) | 147.3<br>(75.0–289.1) | 252.8<br>(132.6–482.0) | 174.2<br>(64.0–474.5) |

a. Defined as a post-vaccination titre at least four-fold higher than the baseline titre.
b. P-value between Day 215 and Day 243 values.

By the time of this follow up 6 months after the second dose, these responses had declined, but neutralising antibodies persisted above baseline with GMTs on Day 208 of 23·9 (95% CI 14·0– 40·6) by PRNT and 54.6 (41·7–71·5) by MNT; both values were significantly lower than the Day 56 values (both p < 0·0001). The majority of the 175 participants still had neutralising antibody titres above baseline: 133 (76%) by PRNT and 155 (87%) by MNT. Following the decline in titres, we were able to estimate decay rate constants (k) of 0·01 and 0.02 based on PRNT_50_ and MNT_50_ titres, respectively (*Supplementary figure 1*). Based on the exponential decay model, the estimated half-life of neutralising antibody titres of all participants was predicted to be 45·6 days for PRNT_50_ and 24·3 days for MNT_50_.

On Day 243, four weeks after the booster vaccination administered on Day 215, there were statistically significant increases in neutralising GMTs, ∼30-fold when measured by PRNT (746 [95% CI: 515–1081], p < 0·0001) and ∼10-fold by MNT (641 [537–765], p < 0·0001). Seroconversion rates increased to 98·7% and 100%, respectively. Higher levels of neutralising antibodies were observed with both assays than those observed after the two doses primary series (**Table 2**). GMTs and seroconversion rates also increased in the placebo group, but to a lesser extent than the booster group — GMTs increased ∼4-fold to 101 (43·6–233) and 359 (267–483) for PRNT and MNT, respectively, with corresponding increases in seroconversion rates to 79·8% and 92·9%. The increase on Day 243 following booster vaccination was significantly higher than in the placebo group when assessed by either PRNT or MNT assays (p < 0·005). Final MNT_50_ GMTs in both groups were statistically higher, p < 0·0001 for the booster arm and p < 0·005 for the placebo arm, than the GMT of 103 (95% CI 50·3–212) observed for the BEI reference sera.

### SARS-CoV-2 variants

Neutralising antibody GMTs against the SARS-CoV-2 variants (Alpha, Beta, Delta and Delta plus) were assessed by PRNT on Days 208 and 243 in the vaccine group; no samples were analysed in placebo recipients. At Day 243, the GMT against the BBV152 strain (D614G) in this group was 632 PRNT_50_ (95% CI: 380–1052). As shown in **Table 2**, GMTs against Alpha, Beta, Delta and Delta plus variants were 338 (188–607), 147 (75·0–289), 253 (1336–482) and 174 (64·0–475) following 10·9-, 161·0-, 264·7-, 32·6- and 174·2-fold rises from Day 208, respectively. The GMT ratio between the BBV152 strain-D614G vs the Delta variant was 2·5 (2·87–2·18).

### IgG binding antibodies by ELISA

The GMT of specific binding IgG antibody titres against S-protein on Day 56 was 9,542 (8,246– 11,041). At Day 215, titres remained above baseline, but the GMT had declined to 4061 (292– 5,647) and 5,115 (3,766–6,948) in the booster and placebo groups, respectively. These GMTs were increased at Day 243 to 11,119 (8,689–14,229) and 7,109 (5,316–9,505) after booster and placebo, respectively (*Supplementary Table 2*). In a similar pattern, the GMTs of anti-RBD and anti-nucleocapsid IgG antibodies in the booster and placebo groups declined from Day 56 to Day 215 but increased at Day 243 following both booster and placebo, respectively (*Supplementary Table 2*). These changes illustrate the responses observed as neutralising titres to booster vaccination and natural exposure in the placebo group were also present as changes in IgG antibodies.

Th1 and Th2 dependent immunoglobulin subclasses (IgG1 and IgG4, respectively) measured by ELISA on Days 215 and 243 showed Th1-biased response in both vaccine and placebo groups, as reported earlier [6,8]. On Day 215, the Th1:Th2 index in vaccine groups was 10·0, which increased to 16·0 on Day 243 (*Supplementary Figure 3*).

### Cell-mediated immunity

On Day 208, six months after the second dose, the median IFN-γ release obtained from SARS-CoV-2 stimulated T cells of vaccinated subjects was 148·8 ng/ml, which is statistically significantly (p < 0·0001) higher than in the corresponding unstimulated cells, 26·6 ng/ml (*Supplementary figure* 2). This indicates the persistence of T cell responses up to 6 months after the second dose. Vaccine-induced SARS-CoV-2 recall responses were demonstrated by the presence of SARS-CoV-2-specific CD4^+^ or CD8^+^ central (CCR7^+^CD45RA^−^, T_CM_), effector (CCR7^−^CD45RA^−^, T_EM_) and terminally differentiated effector memory cells that re-express CD45RA (CCR7^−^CD45RA^+^, T_EMRA_) T cell population (*Supplementary Table 3*). On day 215, among the SARS-CoV-2-specific CD4^+^ T cell population, the proportion of T_CM_ were 34·6% and 34·8% in vaccine and placebo groups, respectively, and the proportions of T_EM_ cells were 42·7% and 40·8%. In contrast, proportions of CD4^+^ CCR7^−^ CD45RA^+^ (T_EMRA_) cells are minimal, 0·2% and 0·0% (**Figure 4A & 4B**) in both groups. In contrast, among the SARS-CoV-2-specific CD8^+^ T cell population at Day 215, the proportions of T_EMRA_ cells were 26·9% and 13·7% in vaccine and placebo groups, while the proportions of T_CM_ and T_EM_ cells were minimal (**Figure 4C & 4D**). On Day 243, vaccine-induced SARS-CoV-2-specific CD4^+^ or CD8^+^ recall memory responses were similar to Day 215 in all the memory phenotypes tested. Collectively, these results demonstrate a phenotypic profile of antigen specific CD4^+^ and CD8^+^ T cells associated with protective immunity to SARS-CoV-2 infection with a good antigen recall response.

**Figure 4.**
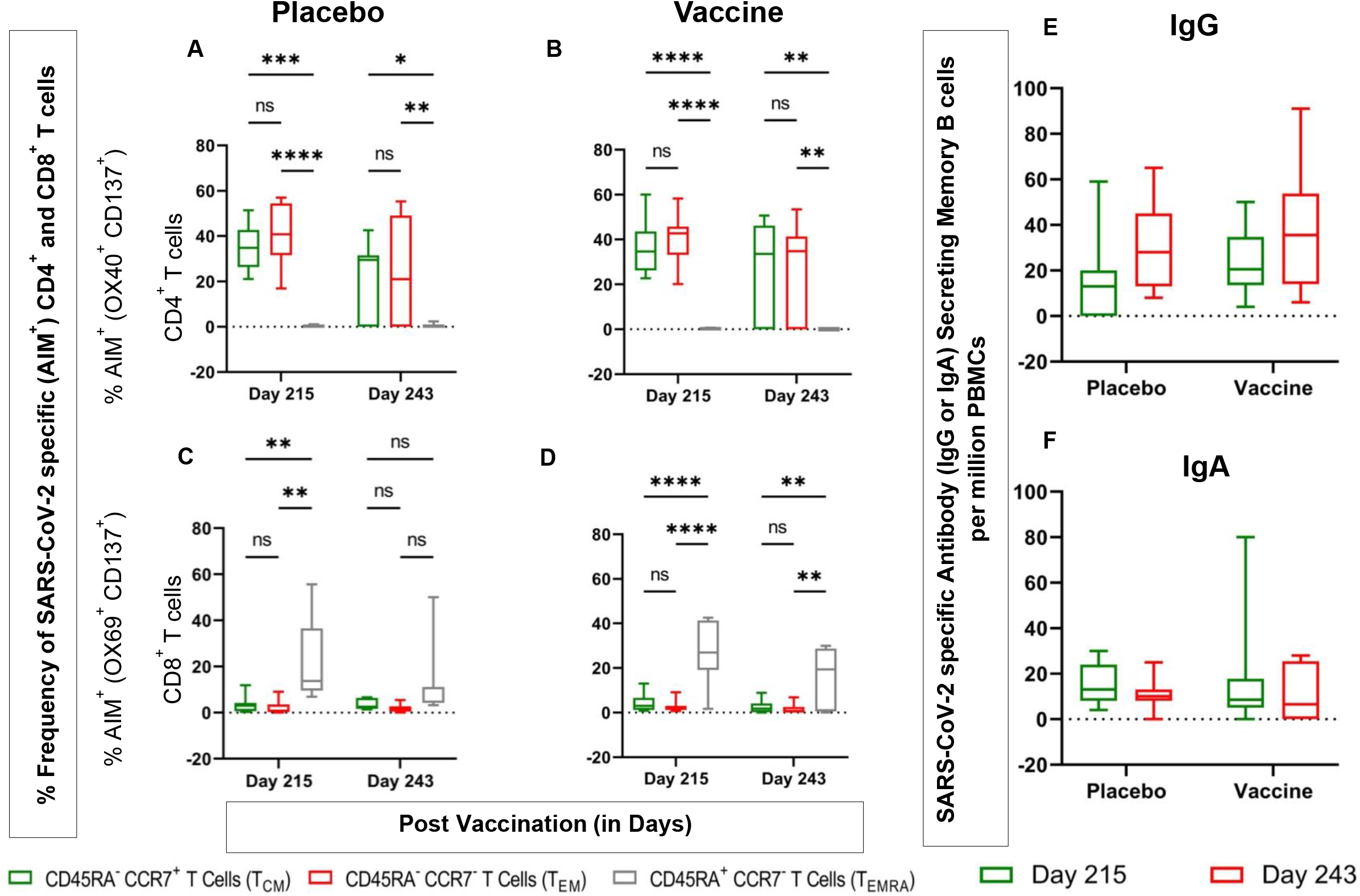
Persistence of SARS-CoV-2 Cell-Mediated Responses **Figure 4** Box and Whisker Plot displays the SARS-CoV-2-specific T & B cell memory response before and after 3rd dose of BBV152. PBMC samples were collected from 15 participants (n=08 and n=07 collected from two-dose BBV152 recipients) on day 215 (before 3rd dose) and 243 (28 days, after 3rd dose). Boxes indicate upper and lower quartiles. The line within the box indicates median and lines extending from the boxes (whiskers) indicating variability outside the upper and lower quartiles Figure 4A, B, C & D represents SARS-CoV-2-specific T cell memory responses were evaluated by performing an activation-induced marker (AIM) assay. PBMCs were stimulated with a cocktail of SARS-CoV-2 peptide pool (S, M, and N) (1μg/mL) for 24 hours. The frequency of AIM+ (CD137+OX40+) cells and AIM+ (CD137+CD69+) cells among CD4+ (A & B) and CD8+ T cells (C & D) were gated to analyse the frequency of antigen-specific central, effector and TEMRA populations. (A & B) Frequency of antigen specific TCM, TEM and TEMRA population among CD4+ T cells observed on days 215 and 243 in placebo and vaccine groups, respectively. (C & D) Frequency of antigen-specific TCM, TEM and TEMRA population among CD8+ T cells observed on days 215 and 243 in placebo and Vaccine groups, respectively. Figure E & F represents vaccine-induced antigen-specific antibody (IgG & IgA) secreting memory B-cell responses performed by ELISpot assay. PBMCs collected from vaccinated subjects on day 215 were pre-activated or pre-stimulated with polyclonal expansion using Poly B stimulant for 4days. Unstimulated cells or cells without pre-activation were used as negative controls. Activated PVDF ELISpot plates were coated with the whole virion inactivated SARS-CoV-2 antigen, and the cells (with or without pre-activation) were added to the plate overnight. Antibody-secreting cells (ASC) were detected by incubating the plate with anti-human IgG (biotin) and anti-human IgA (FITC) antibody followed by streptavidin–ALP and FITC-HRP, respectively. Y-axis shows antigen-specific memory ASC B cells per million PBMC.

Further, persistence of long-lived memory B cells, demonstrated by the detectable levels of SARS-CoV-2 specific IgG and IgA secreting B cells from both vaccine and placebo groups (**Figure 4E & F**), on Days 215 and 243. On day 215, the number of SARS-CoV-2 specific antibody (IgG) secreting memory B cells (MBC) per 10^6^ PBMCs was 20·5 (IQR 13·5–34·8) and 13·0 (0·0–20·0) in vaccine and placebo groups, respectively. The number of antigen-specific IgG secreting memory B cell (MBC) responses increased on Day 243 to 35·5 (14·0–53·8) and 28·0 (13·0–45·0) in vaccine and placebo groups (**Figure 4E**). On Day 215, the number of SARS-CoV-2 specific antibody (IgA) secreting memory B cell (MBC) per 10^6^ PBMCs were 8·5 (5·0– 17·8) and 13·0 (8·0–24·0) in vaccine and placebo groups, respectively. These MBC responses show small reductions on Day 243 to 6·5 (0·0–25·5) and 10·0 (8·0–13·0) in vaccine and placebo groups, respectively (**Figure 4F**).

#### Reactogenicity

After dose 3, there were 8 solicited adverse events in the vaccine groups and 5 in the placebo group. All 8 in the vaccine group were local reactions at the injection site, 5 (5·4%) cases of pain, 2 (2·1%) of itching and one case of redness (1·0%). In the placebo groups, the 5 reports consisted of 2 (2·1%) cases of injection site pain, 2 (2·1%) of fever and 1 (1·0%) instance of headache (*Supplementary Table 3*). Most of these adverse events were mild and resolved within 24 hours of onset. No unsolicited adverse events or symptomatic SARS-CoV-2 infections were reported to investigators through telephone follow-up or site study visits between Day 0 and the scheduled visit on Day 243. Additional telephone calls were conducted to ensure complete documentation of any breakthrough infection, but routine RT-PCR for SARS-CoV-2 was not done. No serious adverse events, including hospitalisation and death, were reported through Day 243.

## Discussion

We show that two doses of BBV152 (6 µg with Algel-IMDG) elicited durable neutralising antibody responses until 6 months after the second dose, together with persistent T cell and B cell responses. There was a decline of antibody levels after the second dose of vaccination, in concordance with earlier reported literature [10,11], but more than 75% of all participants followed up 6 months post the second dose still had detectable neutralising antibody responses to the homologous vaccine SARS-CoV-2 strain (D614G). Further, 28 days after a booster dose of BBV152, there was a marked increase in titres to higher levels than those achieved after the two-dose primary series. When assessed against heterologous strains representing the predominant Variants of Concern, humoral immunogenicity increased (19- to 97-fold) towards both homologous and heterologous SARS-CoV-2 variants. Neutralising antibody titres were comparable to an internationally accepted reference panel.

Booster vaccination was well tolerated with few adverse events, the most frequent being mild and transient pain and itching at the injection site. No severe or life-threatening solicited adverse events were reported, and no significant safety differences were observed between BBV152 and control groups; nor were there any safety concerns raised when reactogenicity of the booster was compared with the primary vaccination series. Although the study was not powered to compare such differences, after any dose, the combined incidence rate of local and systemic adverse events is noticeably better than the rates for other SARS-CoV-2 vaccine platform candidates [12–15] and comparable to rates observed for other inactivated SARS-CoV-2 vaccine candidates [16,17]. However, other vaccine studies enrolled different populations and employed varying approaches to measure adverse events.

As previously reported, BBV152-induced antibodies from previously sero-naive (negative for SARS-CoV-2 serology and nucleic acid testing before vaccination) showed no significant decrease in neutralisation activity against the Alpha (B.1.1.7) variant but demonstrate marginal reductions in neutralisation activity, by 2-, 2-, 3-, and 2·7-fold, respectively, against the B.1.1.28, B.1.617.1, B.1.351 (Gamma), and B.1.617.2 (Delta) variants [18–21]. Here, we report lower neutralisation activity against wild type and VOC SARS-CoV-2 strains on Day 208, 6 months after the second vaccination, but following the third dose of BBV152, serum neutralising antibody titres demonstrated 33-fold and 265-fold increases against D614G (the BBV152 vaccine strain) and the Delta variant, respectively. The PRNT GMT ratio for D614G to Delta was 2·5 (1·9–2·2), indicating a comparable neutralisation profile after three doses of BBV152. Neutralisation assays against Omicron are being conducted.

Durable and persisting immune memory against SARS-CoV-2 has been noted after natural infection [22–24]. We found a pronounced SARS-CoV-2-specific T cell response to BBV152, with a majority of CD4^+^ T central (CD4^+^ T_CM_) and effector phenotype (CD4^+^ T_EM_), including distinct CD8^+^ T_EMRA_ phenotype, before and after the booster dose with a good antigen memory recall response. This may allow BBV152 to confer long term protective efficacy against SARS-CoV-2 variants. Vikkuti et al [25] have shown the potential of BBV152 to induce spike and nucleospecific circulating Tfh (cTfh) cells—Tfh1 (CXCR3+CCR6-), Tfh2 (CXCR3-CCR6-) or Tfh17 (CXCR3-CCR6+)—that help in B cell production. Distinct CD8^+^ T_EMRA_ phenotype induced by BBV152 could be attributed to the memory response against conserved nucleoprotein, indicating the nucleoprotein as a potential target for SARS-CoV-2 vaccines. As current VOCs show major mutations in the viral spike protein, if BBV152 vaccine induces immune memory against conserved nucleoprotein it may provide an additional advantage to protect against immune escape variants. In addition, a small increase in the levels of memory B cells after the booster was correlated with an increase in the neutralisation potency against both homologous and heterologous strains supporting vaccine-induced memory B cells playing a role in protection against circulating SARS-CoV-2 variants.

An unexpected observation was the small increase in neutralising GMTs on Day 243 compared with Day 215 in those who received a placebo rather than a booster dose. This is most likely to be due to natural infection as this booster study was conducted during the second wave of COVID-19 progression in India, which peaked between March 29, 2021 and July 6, 2021, dominated by the Delta (B.1.617.2) SARS-CoV-2 variant. There were also increases in SARS-CoV-2-specific IgG binding antibodies in some individuals, further suggesting that these individuals had been naturally infected. However, none of the 380 participants who enrolled in the parent Phase 2 study reported any of the adverse events expected to be associated with COVID-19, nor deaths or hospitalisation, through to the end of this booster study.

It is interesting that despite the dominance of the Delta variant during this period, there were cases of COVID-19 detected in the study population. In an efficacy trial, BBV152 displayed 65·2% (95% CI: 33·1–83·0) protection against the SARS-CoV-2 Delta VoC [8]. Real world effectiveness of 50% (95% CI 33–62) and 57% (21–76) after two doses administered at least 14 and 42 days apart was demonstrated against Delta in a cohort of healthcare workers known to be exposed to higher infectious pressure [26]. BBV152 has been shown to produce broad specific cell-mediated responses towards several VOCs [20]. Collectively, these results suggest BBV152 induced T cell and B cell memory responses to protect against infection, although the SARS-CoV-2-specific antibody responses show a decline. There is a report in the literature that SARS-CoV-2 specific memory T cell responses have been reported to remain detectable in convalescent individuals up to 10 months after infection [23]. A recent report demonstrates that administration of a third dose of CoronaVac six months after a second dose is effective in recalling SARS-CoV-2-specific immune responses, with a marked increase in antibody levels [27]. However, unlike ours, that study did not report on persistence of cell-mediated responses at 6 months after the second dose.

Our results do not permit efficacy assessments, although we have demonstrated the efficacy of BBV152 in a larger study [8]. The present study enrolled a limited number of participants aged from ≥12 to ≤ 65 years, and further studies are required to establish the effect of a booster dose in the elderly or immunocompromised. An ongoing longitudinal follow-up of additional post-vaccination visits (months 9 and 12) will be important to understand the durability of immune responses. However, this study is the first double blind, randomised, controlled trial to evaluate both humoral and cell-mediated responses of a SARS-CoV-2 booster dose vaccine. In the parent study, six months after the second dose of BBV152, we observed durable neutralising antibody responses that were significantly higher than international reference serum (Source: BEI resources, USA) (Catalog No. NR-53874/Lot No. 70039764).

We have previously reported an interim efficacy of 93% against severe COVID-19 and 65% against any severity of disease due to the Delta variant (median follow-up of 4.7 months after dose 1). This report contains data on the humoral and cell-mediated response from a booster dose of BBV152. No clinical endpoints were evaluated, and the level of neutralising antibodies after the third dose cannot be used to infer any level of protection, although there is data to suggest a correlation between neutralising antibody titres to protection [28]. The Delta variant has been reported to have a shorter incubation period (4 days) compared with the ancestral Wuhan (6 days) [29], so faced with a decline in neutralising antibodies, persistent cell-mediated memory responses may be important to provide durable vaccine efficacy against severe COVID-19 [254]. However, a marked reduction in efficacy against breakthrough infections leading to mild to moderate disease may be expected, and the long-term efficacy of BBV152 against severe COVID-19 is currently being evaluated in a phase 3 study. With the earlier demonstration of 65% efficacy against Delta at 4.7 months median follow-up and a favourable reactogenicity profile of an inactivated vaccine, a broad antibody response to a third booster dose may be advised to ensure robust neutralising antibody titres that prevent breakthrough variant-related mild to moderate disease and infection [30].

In conclusion, the presence of recalled T and B cell responses to SARS-CoV-2 specific antigen stimulation, 6 months after a 2-dose vaccination schedule suggests good immune memory and effectiveness of vaccine against homologous SARS-CoV-2 strain. However, the marked increase in neutralising titres against both homologous and heterologous strains (Alpha, Beta, Delta and Delta plus) with a three dose regimen may necessitate roll out of booster vaccination to provide immune protection against new emerging SARS-CoV-2 variants.

## Data Availability

Individual participant (de-identified) data will be made available when the trial is complete upon a direct request to the corresponding author with an appropriate research proposal. After consideration and the approval of such a proposal data will be shared through a secure online platform.

## Acknowledgements

We would like to sincerely thank the principal and co-principal investigators, study coordinators and healthcare workers involved in this study. We also acknowledge the contributions of Drs. Shashi Kanth Muni, Jagadish Kumar, Vinay Aileni, and Ms. Sandya Rani, Aparna Bathula, Amaravani Pittala of Bharat Biotech to the protocol design and clinical trial monitoring. We thank Drs. Rakeshchandra Meka, for the isolation of PBMCs from whole blood, and Dr. Ramulu Chintala, Spandana Sure and Dr. Krishna Shilpa Palem for their contribution in performing the Immunoglobulin subclass analysis. This vaccine candidate could not have been developed without the efforts of Bharat Biotech’s Manufacturing, Quality Control teams. Keith Veitch (keithveitch communications, Amsterdam, the Netherlands) provided editorial assistance with the final manuscript. All authors would like to express their gratitude to all frontline health care workers during this pandemic.

## Author Contributions

All authors met the International Committee for Medical Editors criteria for authorship. KMV, PY, GS, and BG accessed and verified the data (the CRO was responsible for generating the report). HJ, DR, UP, BG, PY, and GS performed the immunogenicity experiments. SR was the study coordinator and provided invaluable assistance with the protocol design and generation of the interim report. KMV, PS, SR, VS, SP, and RE contributed to the analysis and manuscript preparation, in which PA, SP, NG, and BB of NIV and the ICMR, India participated. KE was responsible for overall supervision of the project and review of the final paper. All principal investigators were involved in the scientific review of this manuscript.

